# Interactions Among Tumor Subtype, PPARγ Expression, and Adipose Proliferation Shape Outcomes in Breast Cancer

**DOI:** 10.1101/2025.07.08.25331111

**Authors:** Aditya Shah, Katie Liu, Ryan Liu, Gautham Ramshankar, Curtis J. Perry, Rachel J. Perry

**Affiliations:** Departments of Cellular & Molecular Physiology, Internal Medicine (Endocrinology), and Comparative Medicine, Yale University, New Haven, CT, United States; Woodbridge Academy Magnet School, Woodbridge, NJ, United States; Cedar Park High School, Cedar Park, TX, United States; Brown University, Providence, RI, United States; Yale Cancer Center, New Haven, CT, United States; Department of Internal Medicine (Hematology/Oncology), Yale University, New Haven, CT, United States

**Keywords:** breast cancer, 18F-FLT, adipose tissue, lipids, PPARγ

## Abstract

Breast cancer progression is influenced by tumor subtype, metabolic environment, and patient factors, including menopausal status and BMI. In this study, we utilize publicly available data to investigate the prognostic relevance of *PPARγ* gene expression, a key regulator of lipid metabolism, and its implications across different subgroups. We also examine the proliferation of adipose tissue in patients with various tumor subtypes and phenotypic cohorts. We analyzed RNA-seq data from 1094 primary breast cancer patients in the TCGA-BRCA cohort, stratifying patients by *PPARγ* expression, menopausal status, and tumor receptor subtype (ER+, HER2-, TNBC) using the UCSC Xena Browser Viewer. Kaplan-Meier analysis revealed that high *PPARγ* expression (≥6.903 FPKM) was significantly associated with both improved overall and disease-specific survival, particularly in premenopausal patients. Complementing this, we analyzed PET-CT scans from 69 breast cancer patients in the ACRIN-6888 clinical trial, focusing on SUV metrics (mean, max, peak) of a cell cycle tracer, 3’-deoxy-3’-[^18^F]-fluorothymidine (^18^F-FLT) in visceral and subcutaneous adipose tissue at the L3/L4 level. Postmenopausal patients had lower visceral SUV_mean_ (0.705 vs 0.776), and patients with ER+ tumors or non-TNBC tumors showed significantly lower SUV_peak_ and SUV_max_ of adipose tissue compared to their counterparts (p < 0.05), indicating metabolic/proliferative reprogramming of adipose tissue based on tumor type. Our study provides a deeper understanding of *PPARγ* as a therapeutic target in breast cancer through lipid metabolism pathways. The changes in adipose tissue proliferation across cohorts demonstrate the strong potential for subtype-specific breast cancer treatment.

## INTRODUCTION

Breast cancer is the most prevalent form of cancer affecting women – it accounts for one-eighth of all cancers diagnosed, with the number of breast cancer diagnoses consistently increasing in recent years [1]. Breast cancer cases are heterogeneous: understanding more about different mechanisms of breast cancer by patient subpopulations will help pave the way for personalized medicine and more effective therapies [2, 3]. Due to this, it is becoming increasingly important to understand the specific metabolic mechanisms that exacerbate the disease – this field was initially explored by Otto Warburg. Warburg pioneered the idea that, due to their need for increased energetic input to continue to proliferate, rapidly dividing cells exhibit a marked increase in glucose uptake [4]. Researchers have continued exploring the central role of glucose metabolism in cancer. However, other metabolic pathways essential for tumor growth have often been overlooked, including lipid metabolism.

Besides the genetic predispositions that contribute to the risk of cancer onset, environmental factors and behaviors can strongly shape and reduce risk. For example, physical exercise reduces fat volumes throughout the body and around tumors, with a specific impact on visceral and subcutaneous fat during cancer development [5]. The typical milieu in which tumors arise is likely also important, as breast cancers develop in lipid-rich microenvironments. Fatty acids can serve as a key energy source for breast cancer cells, whereas other tumors may be predominantly supplied by glucose and amino acids as their main energy sources depend on circulating plasma. This unique environmental association between lipid metabolism and breast cancer makes it particularly interesting to study Peroxisome proliferator-activated receptor gamma (*PPARγ*) gene’s prominence and potential promise as a biomarker in breast cancer [6]. Playing a role in adipocyte differentiation, *PPARγ* is highly expressed in adipose tissue and widely regarded as a master regulator of lipogenesis. Furthermore, reductions in mitochondrial function in cancer have been strongly associated with tumor proliferation – *PPARγ* regulates mitochondrial biogenesis and oxidation by promoting *PCG-1α* (*PPARGC1A*) transcription, thereby enhancing ATP production from fatty acids. The key metabolic hallmark of cancer is a transition from oxidative to glycolytic metabolism [6, 7]. Therefore, it is likely that higher expression of genes that promote oxidative metabolism, such as *PPARγ*, may be protective against tumor progression. The impact of lipid metabolism-related genes, and their interaction with other clinical characteristics, are of particular interest when studying breast cancer, because it is a tumor type that arises in an adipose microenvironment.

Menopausal status is a strong modulator of fat distribution and metabolism, with significance for both breast cancer risk and progression [8]. During menopause, declining hormonal levels (primarily estrogen) shift fat from a dominantly subcutaneous fat distribution to a more visceral fat distribution, increasing the central adiposity, but also altering the metabolic environment. It is particularly interesting to study how patients with different tumor types can be affected during menopause: hormone receptor-positive (HR+) tumors are driven by adipose-derived molecules, making them more common in women with higher adiposity, and perhaps partly as a result also in postmenopausal women [9, 10]. On the other hand, Human Epidermal growth factor Receptor 2 negative (HER2-) tumors are driven more commonly by inflammatory cytokines and metabolic pathways, but can be affected by obesity status and lipid metabolism, making it interesting to study how prognosis can differ in patients with different tumor types [11]. Triple-negative breast cancer (TNBC) tumors are also fueled by immune dysregulation and heightened glycolytic metabolism, and are even more often found in patients with metabolic dysfunction as well [12, 13]. It is important to note that visceral fat is particularly active in releasing inflammatory cytokines which can contribute to tumor progression as well as insulin resistance, amongst other problems [14].

PPARγ is further interesting to study by menopausal status as the reduction in estrogen that occurs with menopause removes a regulatory influence on adipocyte function and lipid metabolism, allowing lipogenesis and fatty acid oxidation pathways, such as those regulated by PPARγ to become more prominent [15, 16]. This makes postmenopausal women particularly susceptible to cancer thriving on lipid metabolism, and developing a further understanding of how PPARγ’s activity is influenced by menopausal status can provide insight into developing targeted therapies for breast cancer prognosis across tumor subtypes. To study these relationships, we utilized a multi-modal approach, performing analysis on Positron Emission Tomography–Computed Tomography (PET-CT) scans from The Cancer Imaging Archive (TCIA), and analyzing the prognostic impact of *PPARγ* expression in the TCGA breast cancer cohort [17–21]. PET-CT imaging is widely used in clinical practice in order to assess metabolic and proliferative activity in tumors and by using radiotracers, such as ^18^F-FLT; using these scans, we are able to quantify proliferation rate in tissue, which is particularly relevant in oncology due to metabolic reprogramming [20, 21]. In this work, we measure the standard uptake values (SUV) of ^18^F-FLT in both visceral and subcutaneous adipose tissue, to understand adipose tissue proliferation in breast cancer patients, and in turn to further contextualize findings from our genomic analyses. After analyzing patient scans and transcriptomics data, we further segment the data by both menopausal status as well as BMI for the PET-CT image analysis, and *PPARγ* expression levels for the Kaplan-Meier survival analysis. This study presents a unique perspective into understanding how lipid metabolism via *PPARγ* gene expression and adipose tissue metabolic activity may impact breast cancer prognosis in patients with varying BMI and menopausal status. These results add to our understanding of breast cancer, paving the way for precision medicine therapies targeting these adipocyte-based pathways.

## RESULTS

### Low *PPAR****γ*** Expression Typically Reduces Survival Across Cohorts

Across our data, by comparing patients with low tumor *PPARγ* expression versus high *PPARγ* expression, we find that typically patients with low expression have worse survivability rates. Across the entire TCGA BRCA cohort, from 1094 patients, we find that patients with high *PPARg* expression (=> 6.903 FPKM) have higher overall survivability rates throughout the entire duration of study (8000+ days), with significance (Fig 1A). The same trend is found by analyzing when studying disease-specific survival, as patients with lower *PPARγ* expression (<6.903 FPKM) have lower survival rates throughout the timeline, however to a lower significance and at a lesser degree (Fig 1B). In the HER2-patient cohort, we again find the same trend to be true with significance, as throughout the entire timeline, patients with high *PPARγ* expression (=>6.866 FPKM), have higher overall survivability rates, indicating that the trend remains similar in patients with HER2 protein in their tumors (Fig 2A). In a contrasting cohort where patients are positive for a receptor type, the ER+ cohort, the same trend holds as patients with high *PPARγ* expression (=>6.803 FPKM), again have higher survivability rates in the entire cohort (Fig 2D). Alternatively, in the triple-negative breast cancer cohort, we find an opposing trend in the long term – initially up to ∼3500 days, the typical trend is seen, where patients with high *PPARγ* expression (=>6.700 FPKM) tend to exhibit higher survivability; however, after 3500 days, patients with high *PPARγ* expression have a trend toward lower survivability rates (Fig 2G), suggesting that high *PPARγ* levels may contribute to long-term outcomes in patients with TNBC. However, these results were not statistically significant and should be considered more suggestive than conclusive.

**Figure 1.**
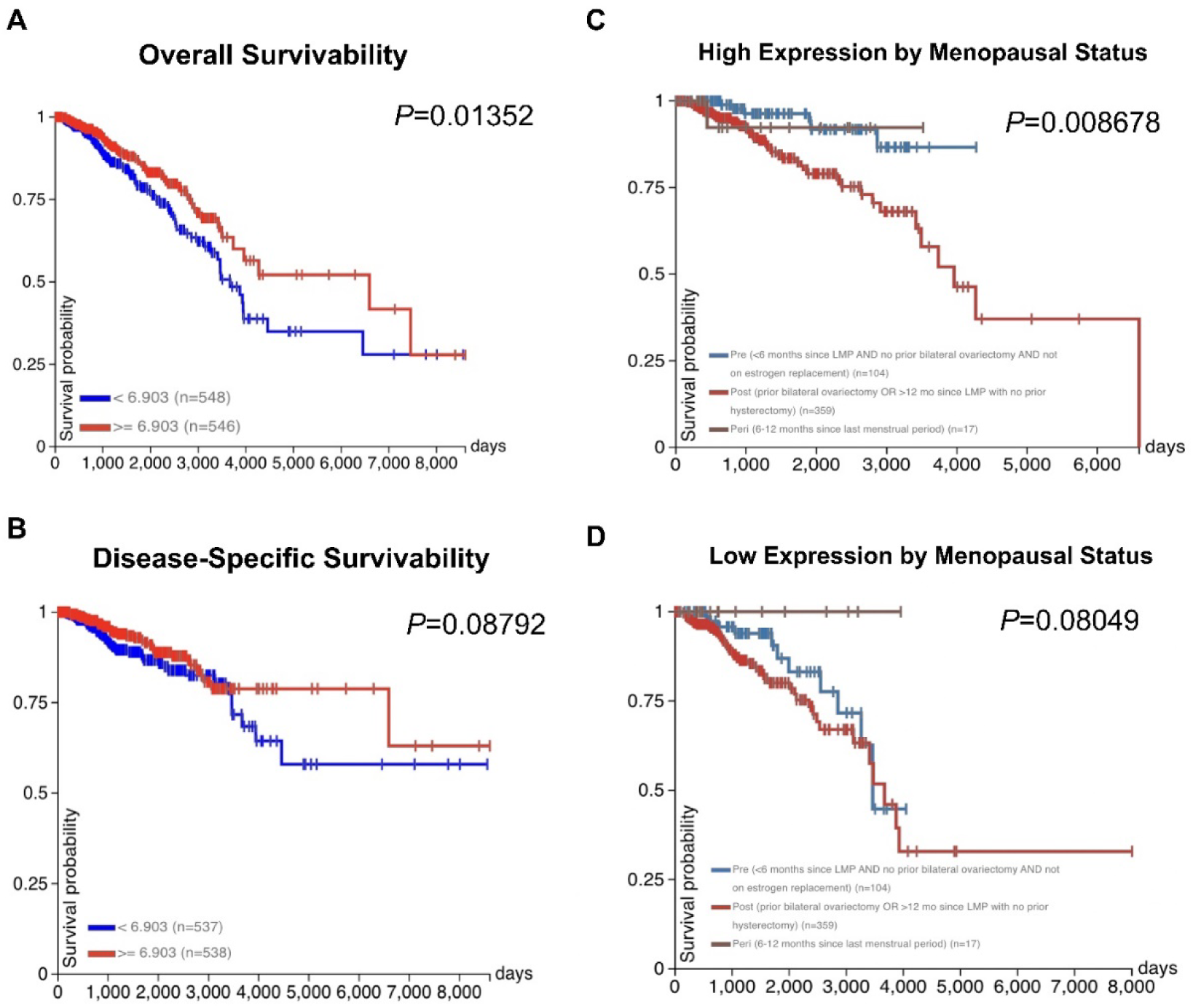
Low *PPARγ* Expression Significantly Reduces Survivability Across TCGA BRCA Patients & Expression Significantly Changes Survival Rate in Pre-Menopausal Patients. **(A)** Overall survivability of patients with low *PPARγ* expression (<6.903 RKPM) in TCGA cohort is consistently lower than those with high expression for 8605 days after initial diagnosis (*P* < 0.01, t-test). **(B)** Disease specific-survivability shows a similar trend, with high *PPARγ* expression corresponding to higher survivability rates throughout, and most significantly after ∼3500 days. **(C)** In patients with high *PPARγ* expression (>= 6.903 RKPM), postmenopausal patients had significantly lower survivability rates, with all postmenopausal patients in the cohort passing away after ∼6500 days. Those with premenopausal or in peri-menopause had significantly better survivability outcomes as shown in the data even ∼4000 days after initial diagnosis, hinting at *PPARγ*’s high expression playing a role in patient survivability during perimenopausal and premenopausal periods. **(D)** In patients with low *PPARγ* expression (<6.903 RKPM), there is not as clear of a relationship between survivability and menopausal status, suggesting low *PPARγ* expression hinders some survivability function that is not specific to menopause, or possibly age.

**Figure 2.**
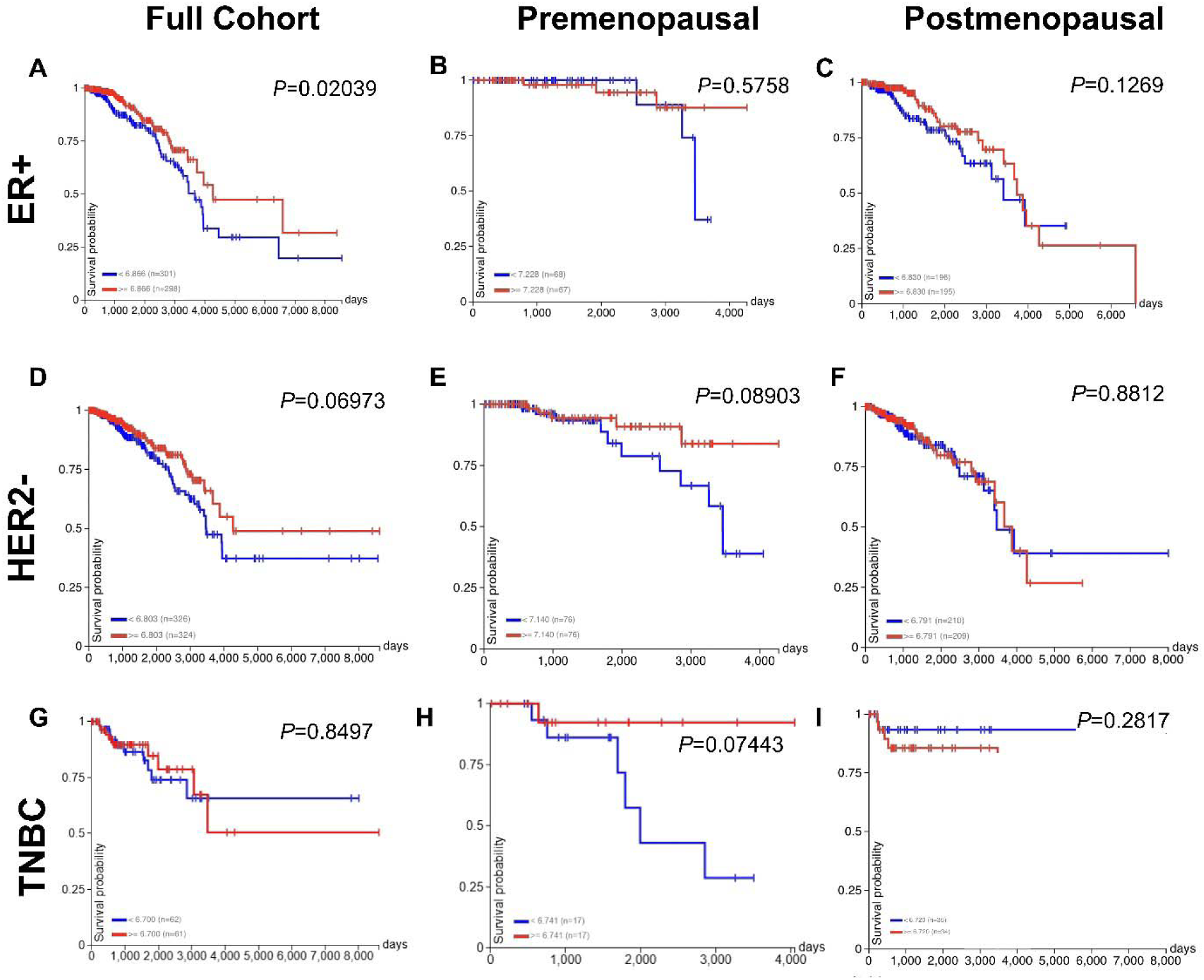
*PPAR γ* Expression Significantly Impacts Survival in ER+ Patients, and TNBC Exhibits an Opposing Pattern. (**A**) Overall survivability in patients with ER+ tumors show the same trend generalized to all primary tumors, those with high *PPARγ* expression (>= 6.866 RKPM), consistently show better survivability rates **(B)** Overall survivability in premenopausal patients with ER+ tumors show the same trend and a steeper decrease in survivability after ∼3000 days for patients with low *PPARγ* gene expression. (**C**) Overall survivability in postmenopausal patients with ER+ shows the same relationship until ∼4000 days. (**D**) Overall survivability in patients with HER2-shows the same trend – those with higher *PPARγ* expression have higher percent of survivability **(E)** Overall survivability in premenopausal patients with HER2-tumors show the same trend as those with primary tumors; while there seems to be no notable difference in the first ∼2000 days, there is a steep difference thereafter. (**F**) Overall survivability in postmenopausal patients with HER2-shows the same relationship through ∼4000 days, and a weak difference thereafter. **(G)** Overall survivability in the TNBC cohort shows a different trend as patients with higher prognosis have a tendency for a lower survival rate after ∼3000 days; however, results are not statistically significant. **(H)** The premenopausal TNBC cohort shows the trend consistent with the full cohort, and different from the TNBC overall data: patients with higher *PPARγ* expression show a marginally significant increase in survival. **(I)** Postmenopausal patients with TNBC tumors show the trend characteristic to TNBC tumors, as patients with high *PPARγ* expression have lower survivability rates across the entire timeline.

### *PPAR****γ*** Expression Correlates with Menopausal Status and Breast Cancer Survival Outcomes

When analyzing data from the entire cohort, and segmenting groups based on menopausal status, we typically find that premenopausal patients have higher survivability rates when compared to postmenopausal patients. This distinction is more clear in high *PPARγ* expression cohorts, where premenopausal patients continuously had higher survival rates, with statistical significance (Fig 1C). While the same trend is true in the low *PPARγ* expression cohort, it is not significant, and even in the long term nearing 3500 days, postmenopausal patients have a higher survivability rate for ∼500 days (Fig 1D). In the premenopausal cohort, across tumor receptor types (HER2-, ER+, TNBC), patients with low *PPARγ* expression have lower survivability rates, especially in the ER+ and TNBC cohorts where there is marginal significance; in the HER2-cohort, there are minimal differences across the two groups (Fig 2B, Fig 2E, Fig 2H). In the postmenopausal group the differences are not as clear, especially in the ER+ group where no differences can be seen until ∼4000 days, where patients with high *PPARγ* expression have lower survivability rates (Fig 2F). The same trend is consistent in the postmenopausal cohorts of patients with HER2- and TNBC tumors, as patients with high *PPARγ* expression have lower survivability rates (Fig 2C, Fig 2I).

### Adipose Tissue SUV Metrics Vary by Menopausal Status and Tumor Receptor Status

Across cohorts, both visceral and subcutaneous adipose tissue SUV_mean_ do not significantly differ between any cohort versus its given alternative (ie. premenopausal vs postmenopausal) (Fig 3A). Similarly, patients who were hormone receptor-positive for a given receptor did not exhibit differences in SUV_mean_ for either VAT or SAT as compared to patients with hormone receptor-negative tumors (Fig 3B-D). On the other hand, SUV_peak_ and SUV_max_ showed significant differences in the TNBC and ER tumor comparisons, and the trend was similar to that found with the SUV_mean_ analyses – patients whose tumors were HR+ had lower SUV values (Fig 4C-D); comparisons of SUV_peak_ and SUV_max_ between menopausal cohorts and HER2 cohorts showed no significant differences (Fig 4A-B).

**Figure 3.**
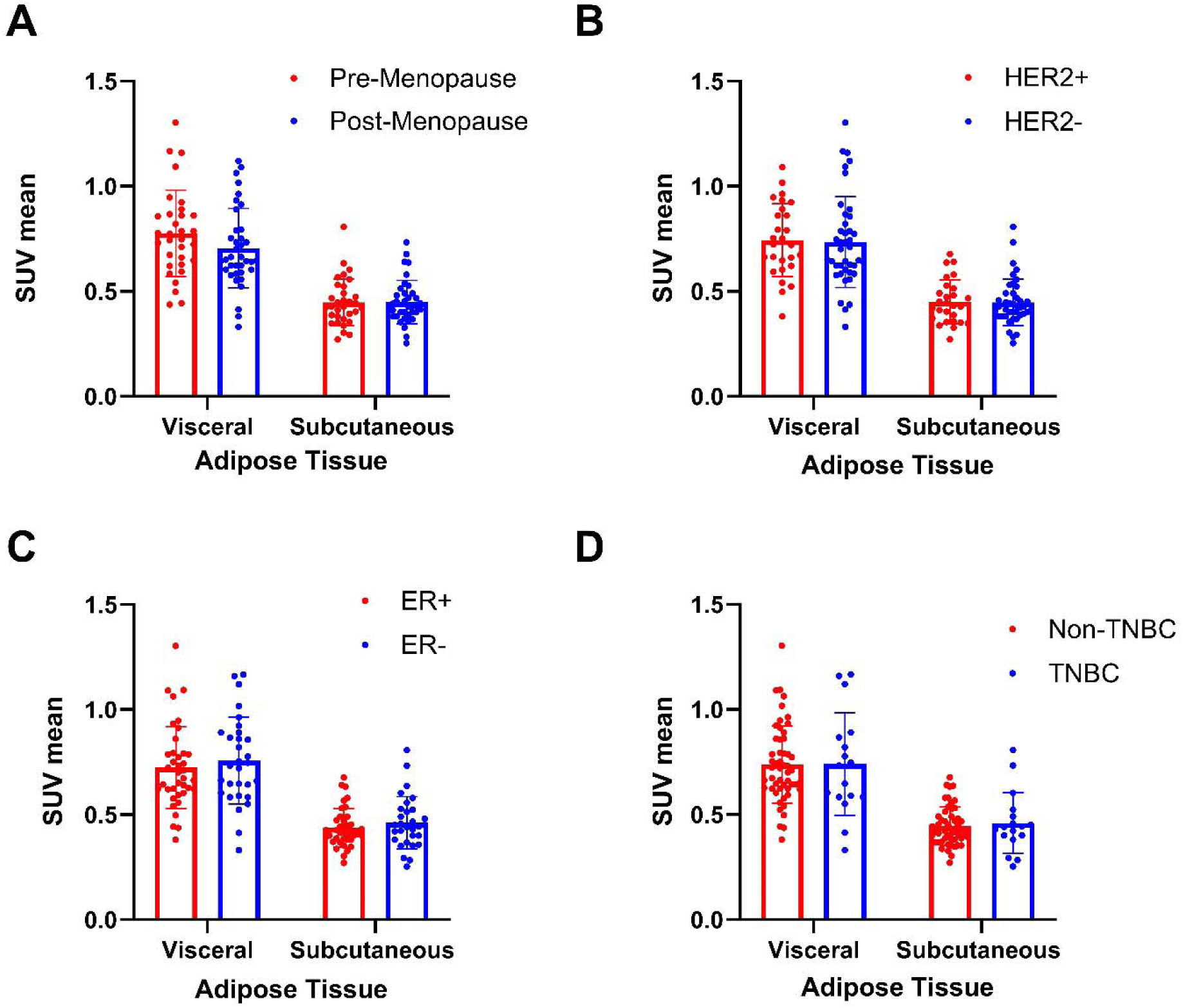
Adipose Tissue SUV_mean_ Remains Consistent Across Tumor Types and Menopausal States in Breast Cancer. **(A)** Visceral adipose tissue SUV_mean_ shows a decrease in post-menopausal (N=37) patients falling from 0.776 to 0.705 in pre-menopausal patients (N=32), while subcutaneous adipose tissue SUV does not show any changes, indicating differential metabolic uptake mechanisms in visceral tissue, data is presented as mean ± SD. **(B)** visceral and subcutaneous adipose tissue SUV_mean_ show very slight decreases in SUV_mean_ in patients with HER2-tumor types (N=41) vs patients with HER2+ tumors (N=27); mean ± SD. **(C)** Patients with ER+ tumors (N=38) have decreased SUV_mean_s of both visceral (0.724 vs 0.757) and subcutaneous adipose tissue (0.436 vs 0.462) versus patients with ER-tumors (N=30), indicating decreased metabolic activity in fat tissue. **(D)** Patients with TNBC tumors (N=17) show a slight increase in both visceral adipose (0.740 vs 0.738) and subcutaneous adipose tissue (0.459 vs 0.444) SUV compared to patients without TNBC tumors, however differences are not statistically different.

**Figure 4.**
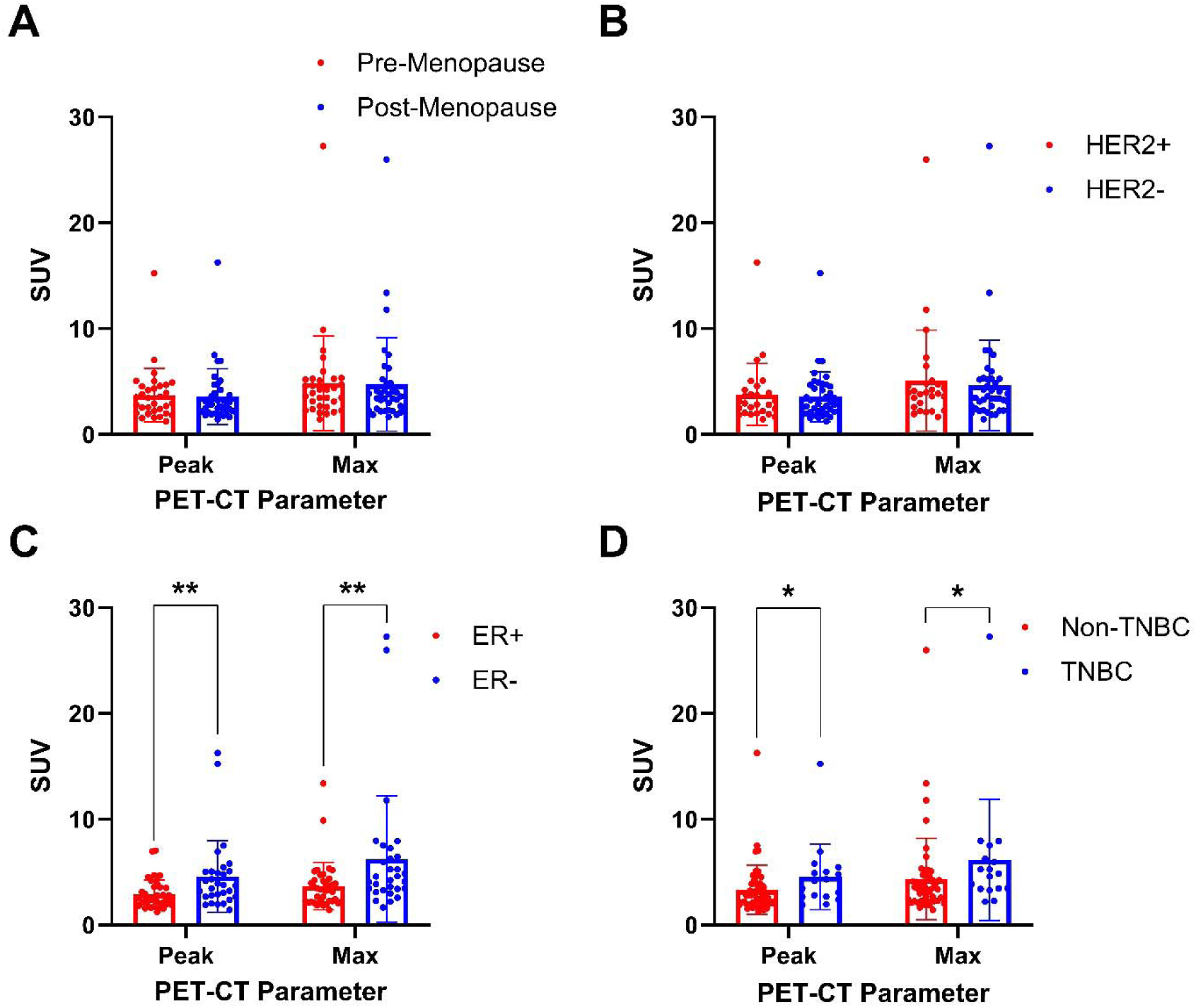
Standard Uptake Value Metrics (g/mL) of adipose tissue differ significantly for ER+ and TNBC tumor types. **(A)** SUV_peak_ and SUV_max_ show no difference based on menopausal status, with a negligible decrease in the postmenopausal cohort for both tissue types. **(B)** HER2-tumors also show a slight decrease in SUV metrics compared to HER2+ tumors, but the differences are not significant. **(C)** ER+ tumors shows significantly lower SUV_peak_ (4.593 vs 2.903) and SUV_max_ (6.245 vs 3.696) in adipose tissue when compared to ER-tumors (*P* < 0.01). **(D)** Patients with TNBC tumors show significantly higher SUV_peak_ (4.566 vs 3.317) and SUV_max_ (6.1617 vs 4.334) in adipose tissue versus patients with any positive receptor tumor types (*P* < 0.05).

### Adipose Tissue SUV_mean_ is Correlated with BMI in Breast Cancer Patients

Across both menopausal subgroups, we evaluated how VAT and SAT tissue metabolic activity, specifically through SUV_mean_, relates to patient BMI. In premenopausal patients, there was no significant correlation between BMI and SUV_mean_ in either visceral adipose tissue VAT or SAT. In VAT, patients displayed no observable trend (r = −0.0772), while in SAT, the correlation was even weaker (r = −0.0258), suggesting minimal metabolic variation by BMI in younger patients (Fig 5A–B).

**Figure 5.**
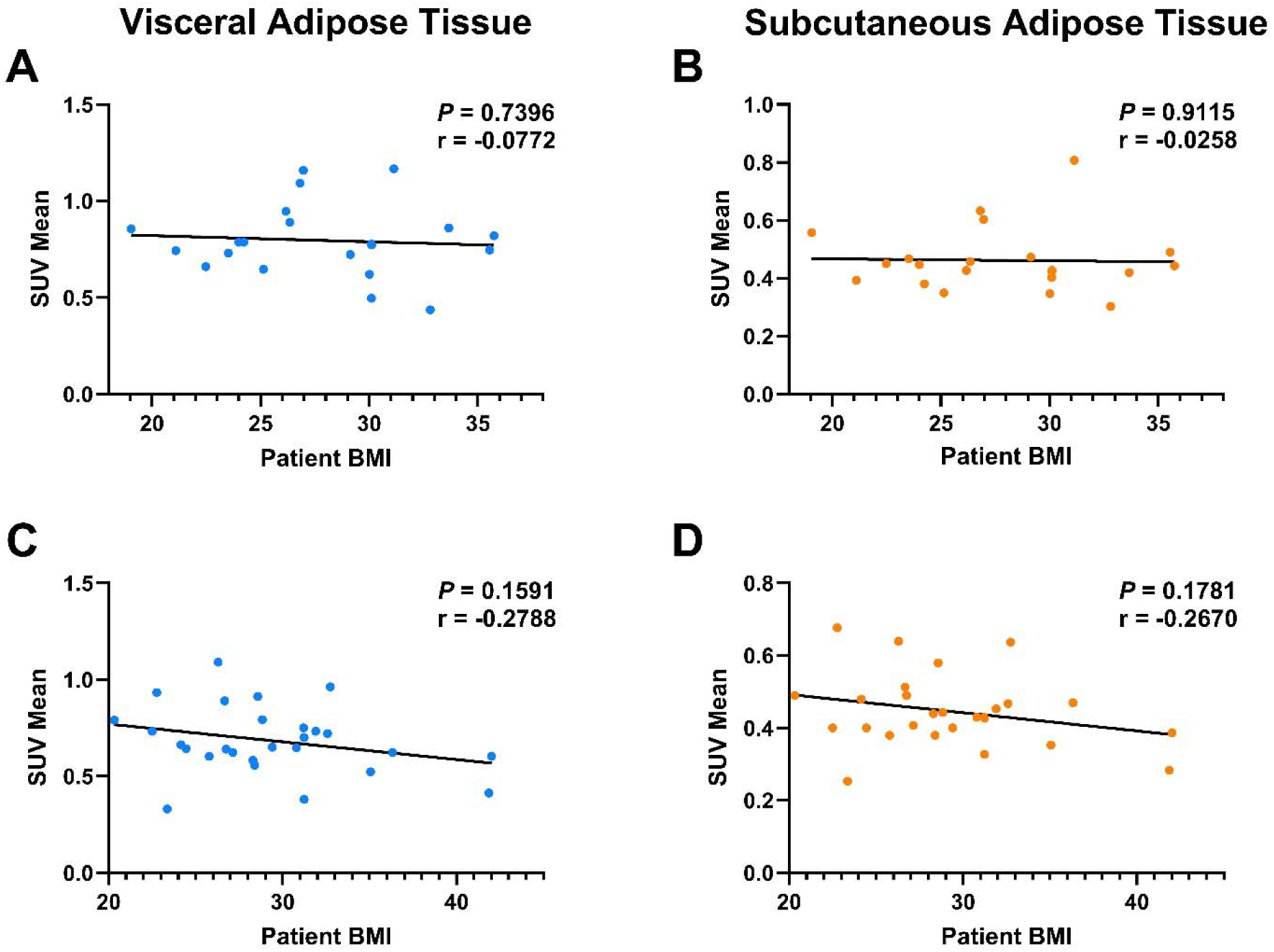
BMI shows stronger correlation with SUV_mean_ in postmenopausal patients. (**A, B**) Premenopausal patients (N=20) show a weak, non-statistically significant negative correlation between BMI and Visceral Adipose Tissue SUV_mean_ (r = −0.077) and an even weaker relationship between BMI and Subcutaneous Adipose Tissue SUV_mean_ (r = −0.0258). (**C, D**) While still not statistically significant, postmenopausal patients (N=28) show a stronger negative correlation between adipose tissue SUV_mean_ and patient BMI for both VAT (r=-0.2788) and SAT (r= −0.2670).

In contrast, postmenopausal patients showed a somewhat stronger trend toward an inverse relationship between BMI and SUV_mean_ in both fat compartments. While not statistically significant, both VAT (r = −0.2788) and SAT (r = −0.2670) trended toward lower metabolic activity in patients with higher BMI (Fig 5C–D). These trends imply that adipose metabolic function may decline with increasing BMI more evidently in older patients. When further stratifying patients by tumor receptor status, we again observe a significant inverse relationship between VAT SUV_mean_ and BMI in patients with ER+ tumors (r = −0.4351), suggesting that ER+ tumors may be particularly influenced by adipose metabolic status (Fig 6C). Patients with HER2-tumors also showed a weak negative correlation (r = −0.1427), though not significant, while in TNBC patients, there was no relationship observed between BMI and VAT SUV_mean_ (r = −0.0742), suggesting the absence of a relationship between visceral metabolic activity and BMI in patients with triple-negative tumors (Fig 6A, E). For SAT, similar trends were seen but to a lesser degree. ER+ patients showed a modest, non-significant negative correlation (r = −0.3194), while HER2- and TNBC groups showed essentially no correlation (Fig 6B, D, F).

**Figure 6.**
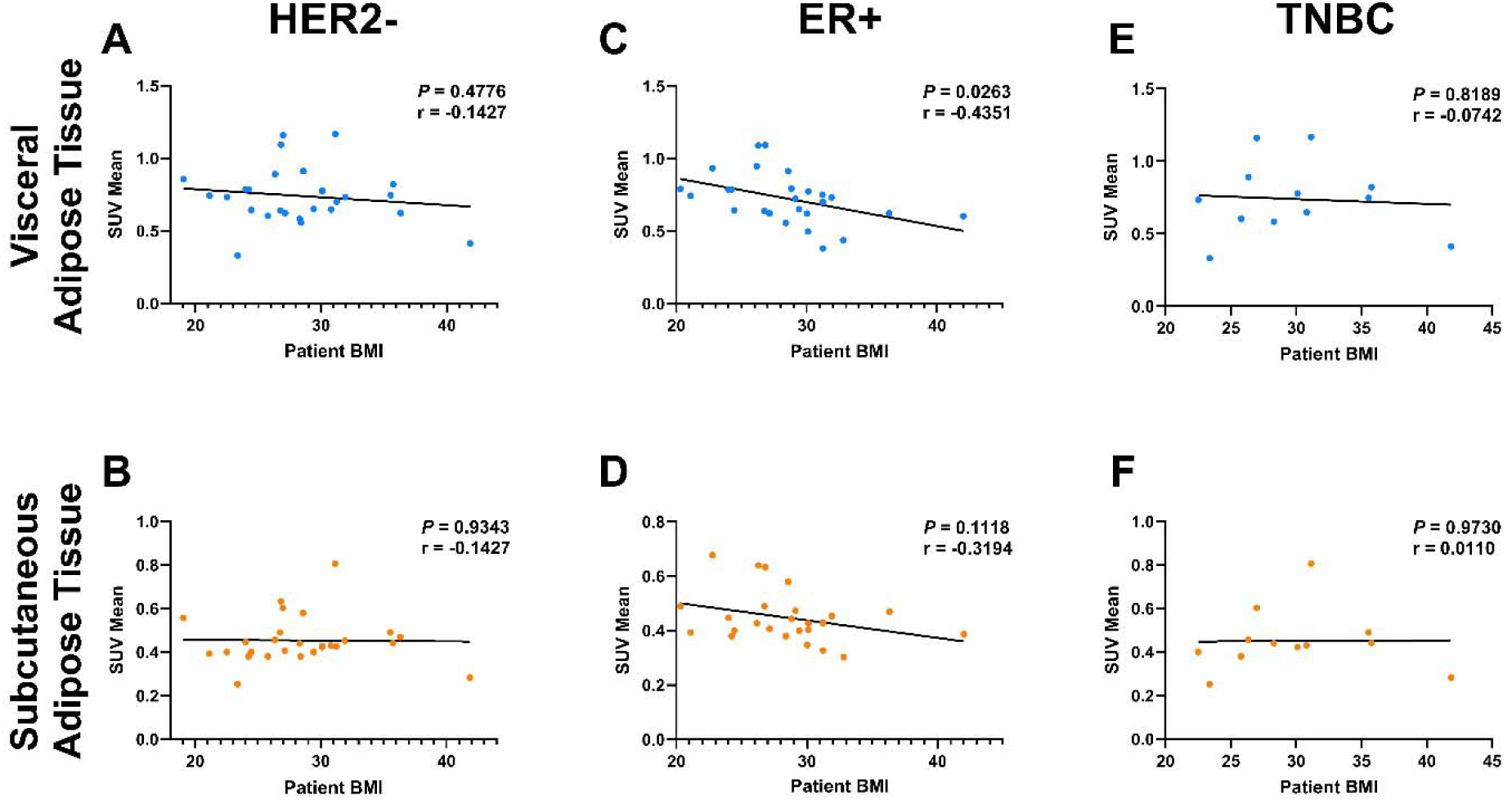
BMI shows a strong correlation with SUV_mean_ of adipose tissue in ER+ tumors; however, the correlation is insignificant in other breast cancer subtypes. (**A, B**) Patients with HER2-tumors show no correlations of adipose tissue SUV_mean_ with BMI. **(C, D)** ER+ tumors show a statistically significant negative relationship between BMI and SUV_mean_, with an r = −0.435 in visceral adipose tissue, and also a negative trend in subcutaneous adipose tissue – both with minimal outliers. **(E, F)** TNBC shows no relationship between BMI and SUV_mean_ in adipose tissue types with r=0.0005 and r=0.0001 in visceral adipose and subcutaneous adipose tissue, respectively.

### Clinical Variables Are Correlated With Adipose Tissue SUV and Tumor Proliferation Metrics

In premenopausal patients, adipose SUV metrics were positively correlated with tumor proliferation markers (mitotic index and Ki-67 index), showcasing a potential link between increased adipose metabolic activity and more proliferative tumors. Notably, VAT SUV_mean_ showed a particularly strong correlation with mitotic index (r = 0.97), and SAT SUV_mean_ also tracked closely with both tumor indices (r > 0.85) (Fig 7A). In contrast, in postmenopausal patients, adipose SUV metrics were inversely correlated with tumor proliferation metrics. VAT SUV_mean_ showed moderate negative associations with both mitotic index (r = −0.44) and Ki-67 (r = −0.60), and SAT SUV followed a similar trend. Furthermore, WBC count was also strongly negatively correlated with VAT SUV_mean_ (r = −0.76), suggesting that higher systemic inflammation may coincide with reduced adipose metabolic activity (Fig 7B). In HER2-patients, adipose SUV metrics were again tightly correlated with each other but showed minimal association with either tumor proliferation indices or WBC count (Fig 7C). However, in ER+ patients, adipose SUV values were quite negatively correlated with both Ki-67 and WBC count (Fig 7D).

**Figure 7.**
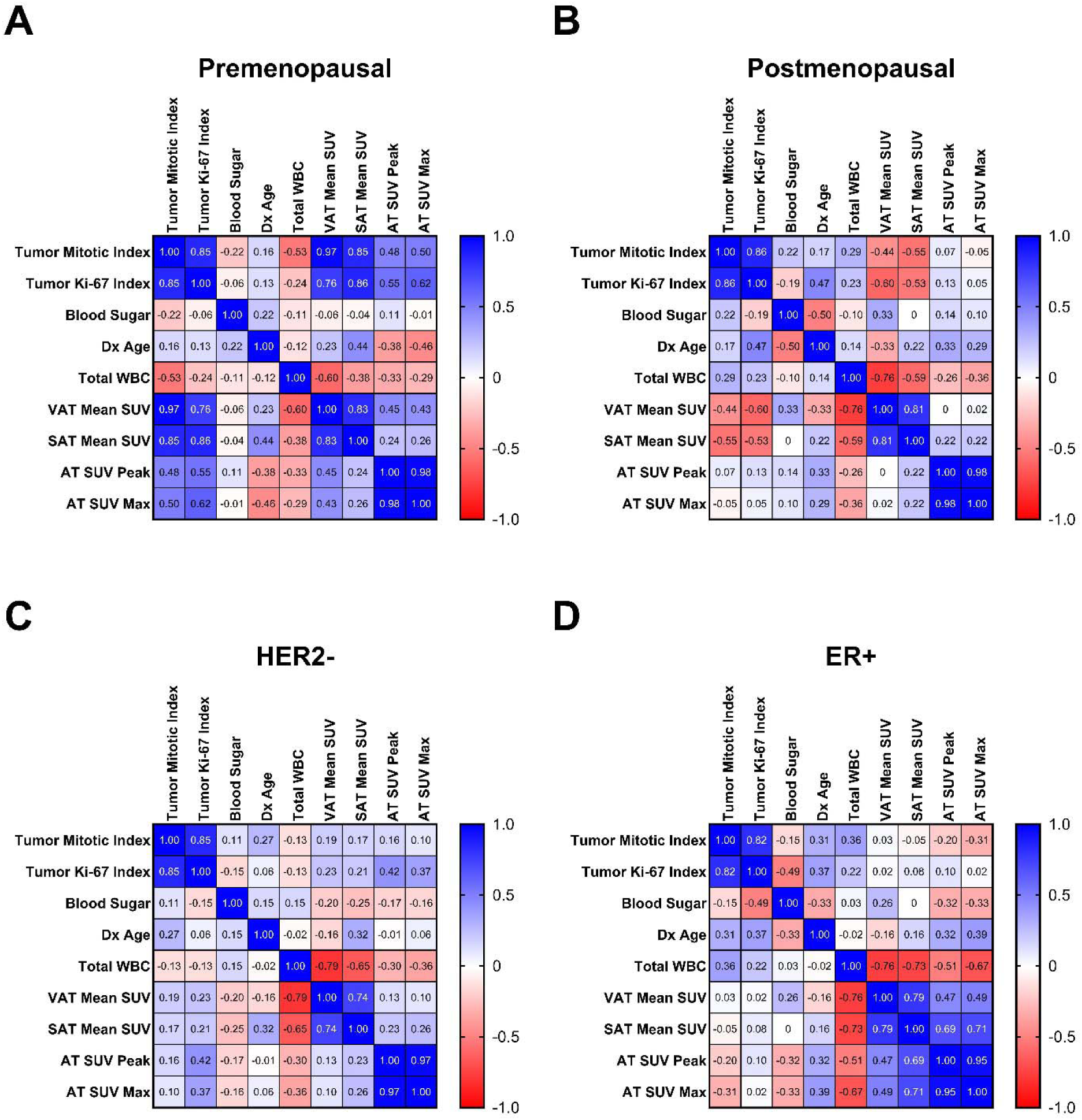
The relationship between SUV_mean_ adipose tissue types and tumor proliferation markers differs between postmenopausal and premenopausal patients, but is similar between patients with HER2- and ER+ tumors. **(A)** In premenopausal patients (N=8), VAT and SAT SUV_mean_ shows strong positive correlations with tumor mitotic and tumor Ki-67 index (r: 0.76 – 0.97), AT SUV_peak_ and SUV_max_ also show positive relationships with tumor proliferation metrics (r: 0.48 – 0.62). **(B)** Postmenopausal patients (N=8) exhibit the inverse relationship, as VAT and SUV_mean_ show negative correlations with tumor mitotic and tumor Ki-67 index (r: −0.60 – −0.44), AT SUV_peak_ and SUV_max_ show no relationship with tumor proliferation metrics. **(C)** In patients with HER2-tumors, clinical variables and proliferation markers show weak relationships across most variables; however, VAT and SAT SUV_mean_ is strongly negatively associated with total WBC (r=-0.79, r=-0.65). VAT and SAT SUV_mean_ are weakly positively correlated with AT SUV_peak_ and SUV_max_. **(D)** Patients with ER+ tumors also have weak correlations between clinical variables and proliferation markers, but show a strong negative correlation of VAT and SUV_mean_ with total WBC as well (r=-0.76, r=-0.73); however, VAT and SAT SUV_mean_ is more strongly positively correlated with AT SUV_peak_ and SUV_max_ (r: 0.49 – 0.71).

## DISCUSSION

In this study, we study PPARγ, a nuclear receptor that plays a central role in adipocyte differentiation, lipid uptake, and anti-inflammatory signaling, making it a compelling candidate for mediating the intersection between systemic metabolism and breast cancer outcomes [6, 22, 23]. Given prior epidemiological and mechanistic evidence linking obesity to a worse breast cancer prognosis in postmenopausal women [8, 24], this study aimed to clarify whether *PPARγ* expression and adipose tissue proliferative activity, as measured by ^18^F-FLT PET-CT SUV metrics, could help explain this association across tumor subtypes and menopausal states. Our findings reinforce that high *PPARγ* expression is broadly associated with improved survival, consistent with its previously documented role to promote oxidative metabolism and suppress pro-inflammatory adipokine signaling in adipose-rich environments such as the breast [16, 25].

Interestingly, this relationship was particularly robust in premenopausal patients and in those with hormone receptor-positive tumors, especially the ER+ subtype (Fig 2). This supports prior literature suggesting that estrogen-responsive tumors are metabolically entwined with adipose-derived cues such as leptin, adiponectin, and free fatty acids—all of which are modulated by PPARL activity [26, 27]. Conversely, in the TNBC subgroup, high *PPARγ* expression was associated with a tendency toward poorer long-term outcomes (Fig 2G–I), a finding that, while not statistically significant, echoes recent work showing that PPARγ may be co-opted by more aggressive tumors to promote immune evasion in nutrient-scarce microenvironments, as was shown recently in hepatocellular carcinoma [28].

We contextualized these transcriptomic findings with PET-CT imaging analysis of ^18^F-FLT uptake in adipose tissue. While average proliferative activity (SUV_mean_) in visceral and subcutaneous fat was relatively consistent across tumor subtypes and menopausal status, SUV_peak_ and SUV_max_ were significantly elevated in patients with TNBC or ER-tumors as compared to those with hormone receptor-positive/HER2-positive or ER+ subtypes, respectively (Fig 4). These higher SUV metrics may reflect enhanced adipose tissue turnover or local proliferative responses, which could be possibly linked to inflammatory or immune cell infiltration – both of which are known to shape the metabolic microenvironment in aggressive breast cancers [29–31]. The consistently (but modestly) lower adipose SUV values in ER+ tumors could reflect PPARγ-mediated metabolic inactivity or lipid storage programs that dampen inflammatory signaling and limit local proliferative stimuli.

Interestingly, our data also showed that the inverse relationship between adiposity (measured via BMI) and adipose SUV_mean_ was only evident in postmenopausal patients, not in premenopausal cohorts (Fig 5). These results align with prior studies, which show that after menopause, declining estrogen levels lead to greater visceral fat accumulation and adipose dysfunction, characterized by reduced mitochondrial activity, increased hypoxia, and elevated secretion of pro-inflammatory cytokines such as IL-6 and TNF-α [32]. PPARγ is known to suppress many of these pathways by promoting fatty acid uptake and oxidation. Still, its expression and efficacy may be attenuated by age-related shifts in the adipose transcriptome [33]. These findings suggest that the menopausal state and adipose tissue composition modulate the prognostic relevance of metabolic genes, such as PPARγ, and may inform stratification strategies for patients.

Additionally, when we examined adipose metabolism in relation to tumor proliferation (via Ki-67 and mitotic index), we found a notable dichotomy. In premenopausal patients, SUV_mean_ in both VAT and SAT positively correlated with tumor proliferation, suggesting metabolic crosstalk or shared growth-promoting signals between fat and tumor tissue (Fig 7A). In contrast, this relationship reversed in postmenopausal patients, where higher SUV_mean_ was associated with lower proliferation (Fig 7B). This may reflect the altered secretory profile of postmenopausal adipose tissue, where hypertrophy and macrophage infiltration lead to the release of lipotoxic factors rather than growth-promoting ones [34]. Alternatively, it may suggest that tumor aggressiveness in older patients becomes decoupled from adipose metabolism and is driven more by tumor factors or immune suppression.

Moreover, across both HER2– and ER+ subgroups, we found strong negative correlations between WBC count and VAT SUV_mean_ (Fig 7C-D), reinforcing a potential link between systemic inflammation and suppressed adipose metabolic function in breast cancer across these subtypes. These findings are consistent with prior work demonstrating that chronic inflammation impairs adipocyte oxidative capacity and to some capacity also promotes insulin resistance— both of which have been implicated in tumor progression and poorer clinical outcomes [34]. PPARγ activation has been shown to reduce WBC-mediated inflammation in metabolic disease and certain cancers, raising the question of whether targeted PPARγ agonism could improve outcomes by restoring adipose function and mitigating paracrine tumor support.

Finally, although BMI is widely used as a proxy for adiposity, it is increasingly clear that visceral adiposity and functional fat metabolism are better predictors of metabolic risk [35] and of cancer risk [36] and mortality [37] than BMI alone. This is underscored by our finding that TNBC patients showed no correlation between BMI and adipose SUV (Fig 6), supporting the idea that BMI may mask underlying metabolic dysregulation in aggressive subtypes. Future studies incorporating visceral fat volume, leptin/adiponectin ratios, and tissue-level lipidomics may provide a more comprehensive understanding of these dynamics. Taken together, our study highlights the utility of integrating transcriptomic and metabolic imaging data to better understand the heterogeneity of breast cancer prognosis. *PPARγ* appears to be most prognostically relevant in ER+ tumors, especially in premenopausal patients, where its metabolic effects are most intact. By contrast, its protective role diminishes with age and may even invert in TNBC, where tumors can exploit lipid metabolism for growth, resulting in an opposite trend. Further studies evaluating pharmacologic PPARγ agonists such as rosiglitazone, particularly in metabolically vulnerable breast cancer subtypes, are warranted to test its translational relevance in breast cancer from our findings, as done in pilot studies in the past [38]. Ultimately, this work reinforces the complex but actionable relationship between systemic metabolism, hormonal status, and tumor behavior in breast cancer.

## CONCLUSION

In summary, our RNA transcriptomic and imaging analyses reveal that high *PPARγ* expression is broadly associated with improved breast cancer survival, particularly in ER+ and premenopausal patients, which we hypothesize is facilitated through enhanced lipid oxidative metabolism and anti-inflammatory regulation (across tumor types). This protective effect diminishes after menopause, where estrogen loss disrupts adipocyte homeostasis and alters PPARγ-driven pathways, as indicated by the less distinct comparisons and further represented by the adipose SUV metrics. Notably, in TNBC, PPARγ may be co-opted to fuel tumor progression, underscoring the subtype-specific metabolic rewiring that is the focus of this study. Together, these data establish *PPARγ* expression as a context-dependent regulator of breast cancer outcome, linking adipose metabolism, tumor hormonal expression, and tumor behavior — supporting its potential as a biomarker and therapeutic target in metabolically vulnerable subgroups, paving the way for more personalized medicine.

## METHODS

### Transcriptomics and TCGA RNA Analysis

After confirming strong expression of *PPARγ* in breast tissue, as shown in the literature and previous studies, we utilized the UCSC Xena Functional Genomics Browser’s Visualization suite to study the prognosis of breast cancer patients in the TCGA Breast Cancer (BRCA) data set, which can be found here: https://xenabrowser.net/datapages/?cohort=TCGA%20Breast%20Cancer%20(BRCA)&removeHub=https%3A%2F%2Fxena.treehouse.gi.ucsc.edu%3A443. The dataset initially contained RNA sequencing data from 1247 patients in column A, which were then filtered for each of the different analyses. Gene expression of *PPARγ* was added to column B, and in column C, sample type was chosen as a phenotypic parameter; after any tumors that did not originate in the breast were removed (ie. non-primary tumors), there were 1101 patients left for analyses, which were then filtered for those with *PPARγ* gene expression data leaving 1094 patients for survivability analysis. These patients were divided at the median for *PPARγ* gene expression in the cohort, which identified low levels of *PPARγ* (< 6.903 FPKM) in 548 patients and high levels of *PPARγ* (>= 6.903 FPKM) in 546 patients. The data from these samples were also examined in order to study disease-specific survivability, which had data from 1075 of the patients and had the same cutoff as described above. For both groups, the survivability was studied for 8605 days (23.5 years) after initial diagnosis, allowing for measures over a long period.

After performing those initial analyses, menopausal status was added as a phenotypic parameter in column D, and the same patients as above were utilized; the menopausal status was either premenopausal, postmenopausal, or perimenopausal. These patients were then further stratified for two analyses, patients with high *PPARγ* expression as described above were selected for one cohort and the other cohort contained data from patients with low *PPARγ* expression – for both of these analyses any patients with an indeterminate menopausal status were removed, which resulted in 491 patients with a high expression that had an available menopausal status. For those with high *PPARγ* expression, 124 were premenopausal, 345 were postmenopausal, and 22 were perimenopausal and for those with low *PPARγ* expression 104 were premenopausal, 259 were postmenopausal, and 17 were perimenopausal. These samples were again visualized with cutoffs set to 6000+ days for both groups after initial diagnosis, to visual long-term survivability relationships.

After performing the analyses non-specifically on all patients with primary tumors, the data were divided by tumor type: ER+ or HER2-tumors were then studied, which was chosen as a phenotypic parameter in column E. After filtering, there were 599 patients with ER+ tumors and 650 patients with HER2-tumors. It is important to note that patients could have both tumor types and that they are not independent of one another. For these initial tumor type-specific analyses, in the ER+ cohort, there were 301 patients with low *PPARγ* expression (< 6.866 FPKM) and there were 298 with high expression (>= 6.866 FPKM), and in the HER2-cohort, there were 326 patients with low expression (< 6.803 FPKM) and 324 with high expression (>= 6.803 FPKM). The cutoffs for *PPARγ* expression dividing patients differ across conditions, as it was determined to be the median of the specific cohort. Patients were then divided by their menopausal status amongst their breast cancer cohort, and it was visualized if *PPARγ* gene expression showed differences between groups of different menopausal statuses. For the ER+ cohort, there were 135 total premenopausal patients, and 68 had low *PPARγ* gene expression (< 7.228 FPKM) and 67 had high expression (>= 7.228 FPKM); there were 389 postmenopausal patients with ER+ tumors, and 196 had low *PPARγ* expression (< 6.630 FPKM) and 195 had high *PPARγ* expression (>= 6.630 FPKM). For the HER2-cohort, there were 152 premenopausal patients, and 76 had low *PPARγ* expression (< 7.140 FPKM) and the 76 others had high expression (>= 7.140 FPKM) and there 419 postmenopausal patients, with 210 of low expression (< 6.791 FPKM) and 209 with high expression (>= 6.791 FPKM). For the TNBC cohort, there were 133 patients overall with available *PPARγ* gene expression data; of these, 62 had low *PPARg* gene expression (< 6.700 FPKM) and 61 had high expression (>= 6.700 FPKM). In the premenopausal subgroup, there were 34 patients: 17 had low expression (< 6.741 FPKM), and 17 had high expression (=> 6.741 FPKM). Among the 69 postmenopausal patients, 35 had low *PPARγ* expression (< 6.720 FPKM), and 34 had high expression (=> 6.720 FPKM).

### Human Patient Image Analyses

In this study, we perform image analysis on PET/CT scans from 69 of the 90 patients from an open-source data repository from The Cancer Imaging Archive (TCIA) - all patients with menopausal status and scans at the L3/L4 level were selected to be part of this study. The dataset can be found here: https://wiki.cancerimagingarchive.net/pages/viewpage.action?pageId=30671268. These patients were part of a clinical trial (ACRIN-6888), all data was de-identified and written consent was provided from the patients during the time of the study. This data was originally approved for public release by the University of Arkansas for Medical Sciences (UAMS) Institutional Review Board (IRB #205568) – all participants provided informed consent for data sharing through TCIA; however, we did not have access to the specific procedures or documentation of the consent process. As the dataset was fully de-identified and its use pre-approved under the original IRB protocol, no additional IRB review was required for our secondary analysis.

Furthermore, 48 out of 69 of the patients studied had their height and weight available, allowing for their BMI to be calculated, and 68 of the patients had their tumor receptor status (ie. HER2 and ER) available, allowing for stratification during the analyses. Each of the patients had scans ranging from their first visit until post-treatment – in order to minimize the effect of the clinical trial itself, and to focus the study on breast cancer cohorts only the first available scans were used for the patient, as treatment for breast cancer can skew overall patient metabolism and tumor proliferation [20]. Overall from the 69 patients selected, 32 were pre-menopausal and 37 were postmenopausal; the BMI of the 48 patients with height and weight available ranged from 19.04 kg/m^2^ to 42.02 kg/m^2^, with 10 healthy (BMI: 18 - 24.9), 18 overweight (BMI: 24.9 - 29.9), and 20 obese patients (BMI: =>30) and the mean of the data 28.64 kg/m^2^ with a standard deviation of 5.03 kg/m^2^. Furthermore, there were 38 patients with ER+ tumors and 41 patients with HER2-tumors - it is important to note that a patient could have both an ER+ and HER2-tumor, as they are not mutually exclusive since the receptors are independent of each other. Patients that were negative for ER, HER2, and PR receptors were selected as part of the triple-negative cohort, which included 17 patients.

Images from each of the patients were uploaded to Fiji ImageJ and using the Brown Fat ROI tool within the PET-CT viewer, we were able to analyze the scans. Since adipose tissue (visceral and subcutaneous) was being studied, we assessed all scans at the L3 or L4 vertebral level, where the tumor is located, and which is the standard landmark for assessing these scans. In particular, to only quantify the standard uptake value of ^18^F-FLT, we set the CT criteria to −190 to −30 Hounsfield Units (HU) to selectively segment adipose tissue, and not other tissue or bone in the surrounding regions. We then drew fixed-volume spheres around the area of interest and were able to calculate the lean body mass-corrected SUV values (including SUV_mean_, SUV_peak_, and SUV_max_), and for each patient, we performed analysis on 3 separate continuous slices in the region. After the SUV values were calculated, patients were separated into their cohorts (ie. by menopause and tumor type) and data were visualized in GraphPad Prism. The dataset also contained patient lab values, including blood sugar, Ki-67 proliferation index, etc – which were used to develop cross-correlation (spearman’s correlation) matrices between these clinical variables and SUV values in adipose tissue, to better understand the interplay across adipose tissue proliferation and these variables. All patients that had all available lab values were used for the subsequent analyses, which included both 8 premenopausal and 8 postmenopausal patients, and also 13 patients with HER2-tumors and 10 patients with ER+ tumors; as there were only 4 patients with TNBC tumors that met the criteria, they were excluded from the analysis due to insufficient power to draw conclusions based on the limited sample size.

## DATA AVAILABILITY

All data analyzed in this manuscript are publicly available at the links in the corresponding parts of the Methods section.

